# Quantitative Assessment of Chest CT Patterns in COVID-19 and Bacterial Pneumonia patients: A Deep Learning Perspective

**DOI:** 10.1101/2020.11.13.20231118

**Authors:** Myeongkyun Kang, Philip Chikontwe, Miguel Luna, Kyung Soo Hong, Jong Geol Jang, Jongsoo Park, Kyeong-Cheol Shin, June Hong Ahn, Sang Hyun Park

## Abstract

As the number of COVID-19 patients has increased worldwide, many efforts have been made to find common patterns in CT images of COVID-19 patients and to confirm the relevance of these patterns against other clinical information. The aim of this paper is to propose a new method that allowed us to find patterns which observed on CTs of patients, and further we use these patterns for disease and severity diagnosis. For the experiment, we performed a retrospective cohort study of 170 confirmed patients with COVID-19 and bacterial pneumonia acquired at Yeungnam University hospital in Daegu, Korea. We extracted lesions inside the lungs from the CT images and classified whether these lesions were from COVID-19 patients or bacterial pneumonia patients by applying a deep learning model. From our experiments, we found 20 patterns that have a major effect on the classification performance of the deep learning model. Crazy-paving was extracted as a major pattern of bacterial pneumonia, while Ground-glass opacities (GGOs) in the peripheral lungs as that of COVID-19. Diffuse GGOs in the central and peripheral lungs was considered to be a key factor for severity classification. The proposed method achieved an accuracy of 91.2% for classifying COVID-19 and bacterial pneumonia with 95% reported for severity classification. Chest CT analysis with constructed lesion clusters revealed well-known COVID-19 CT manifestations comparable to manual CT analysis. Moreover, the constructed patient level histogram with/without radiomics features showed feasibility and improved accuracy for both disease and severity classification with key clinical implications.

## I. INTRODUCTION

The SARS-CoV-2 pandemic virus originated in Wuhan, China in 2019, has spread rapidly to several countries [1]. RT-PCR of viral nucleic acid is regarded as the reference standard for the diagnosis of COVID-19, but chest CT examination is mostly recommended for evaluating severity and treatment efficacy given the primary involvement of the respiratory system. Also, CT imaging can be effective for early screening compared to RT-PCR that has shown low sensitivity for early detection [2]–[4]. Therefore, there is an urgent need for fast and accurate diagnostic tests other than RT-PCR.

There are well-known features of COVID-19 often observed in CT imaging such as GGOs distributed in the peripheral or posterior lungs [5]–[7]. However, these patterns are often limited and contribute to the challenge of distinguishing COVID-19 from other pneumonia types [8], [9]. Furthermore, since COVID-19 CT manifestations are often observed with mixed or subtle radiological differences, accurate description is challenging even when referencing Fleischner Society listed terms [10].

Recent advances in artificial intelligence with deep learning have shown success in the medical imaging community given the robust feature extraction capability of deep networks [11]. Herein, we analyzed chest CT scans from 73 COVID-19 and 97 bacterial pneumonia patients from Daegu, Korea. We propose a deep learning-based framework to create accurate descriptions of CT manifestations related to COVID-19. Specifically, we segmented lung and lesion regions via a segmentation model and then trained a classification model using all lesion patches and assigned them to one of two categories i.e. COVID-19 or bacterial pneumonia. The features extracted by the classification model were clustered into 20 groups via a K-means clustering algorithm [12]. Three experts confirmed that the patterns displayed in each cluster were clinically related to the typical findings of COVID-19.

For each patient, we further constructed a histogram of lesion patches given information of the 20 clusters to obtain a single representative feature vector per 3D CT image alongside radiomics features via concatenation. Finally, the features were employed to train classifiers for (i) distinguishing COVID-19 from bacterial pneumonia patients, and (ii) to assess disease severity (non-severe/severe) cases to confirm the generalization ability of the features. For the first task, the proposed method showed a 3.7% improvement over conventional majority based classification with deep learning, whereas 12.5% was reported for the severity classification task further highlighting the benefit of the proposed feature extraction strategy with significant margins observed across several metrics.

## II. MATERIALS AND METHODS

### A. STUDY DESIGN AND SUBJECT

We performed a retrospective cohort study of CT scans of 73 patients with COVID-19 infection obtained between February 2020 to March 2020, and 97 patients with bacterial pneumonia between March 2012 to February 2014 at Yeungnam University Medical Center, in Daegu, Korea. This study was conducted in accordance with the tenets of the Declaration of Helsinki and was reviewed and approved by the Institutional Review Board of Yeungnam University Hospital (YUH IRB 2020-05-030). The requirement for informed consent was waived due to the retrospective study design. During the study period, all consecutive adult patients (age > 18 years) with SARS-CoV-2 infection admitted to the hospital were eligible for inclusion. SARS-CoV-2 infection was confirmed by real-time RT-PCR assay of nasal and pharyngeal swab samples. Severity was defined as a composite outcome of acute respiratory distress syndrome (ARDS), intensive care unit admission, or death. ARDS was diagnosed according to the Berlin definition [13]. Figure 1 shows the flow chart of data collection, exclusion and splitting ratios applied for training and evaluation.

**FIGURE 1.**
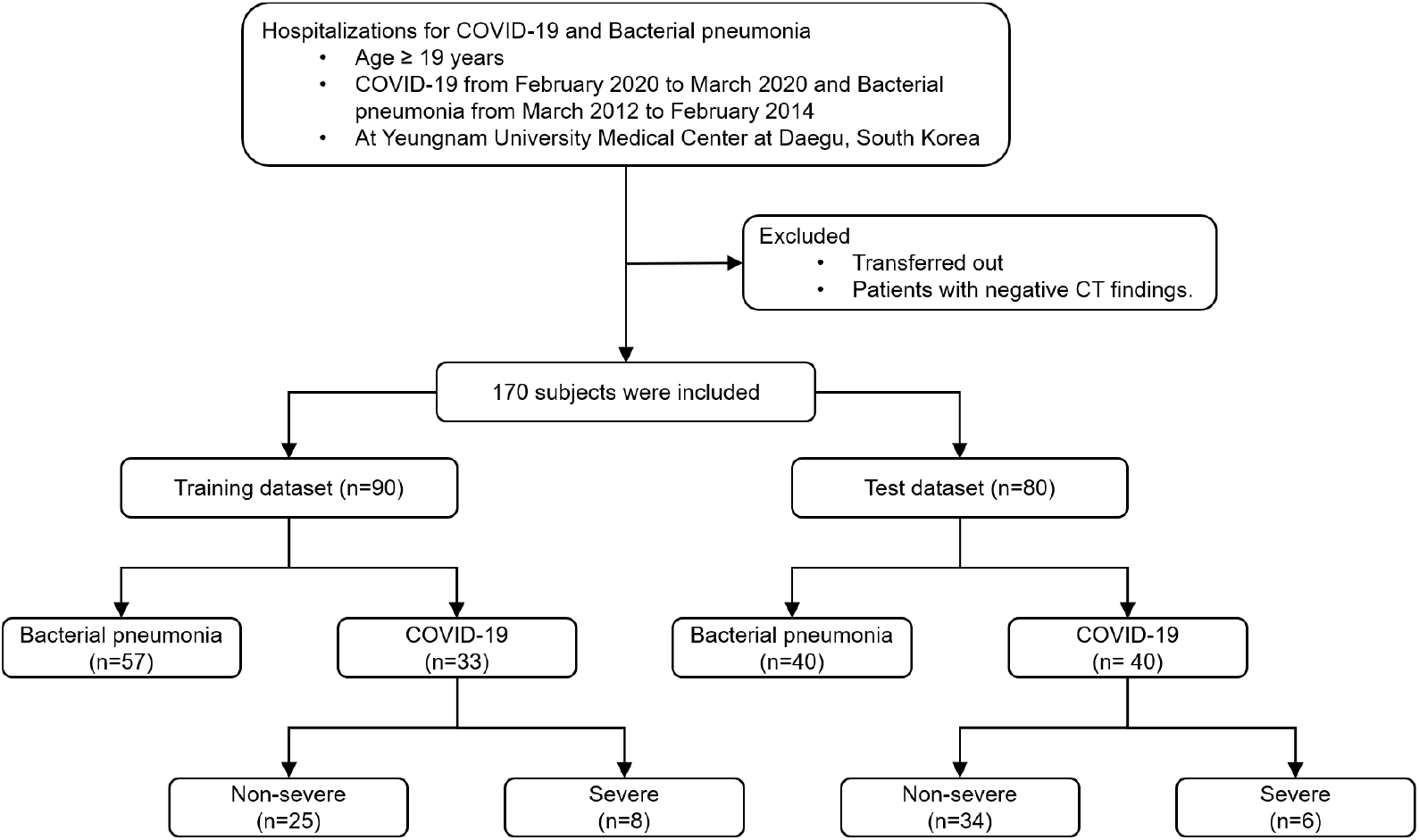
Flow chart.

### B. IMAGING PATTERN ANALYSIS USING DEEP LEARNING

To analyze the COVID-19 manifestations using deep learning, the proposed framework consists of three key modules i.e. (a) lung and lesion segmentation, (b) deep feature extraction, and (c) K-means clustering modules, respectively. Figure 2(a) shows a diagram of the framework.

**FIGURE 2.**
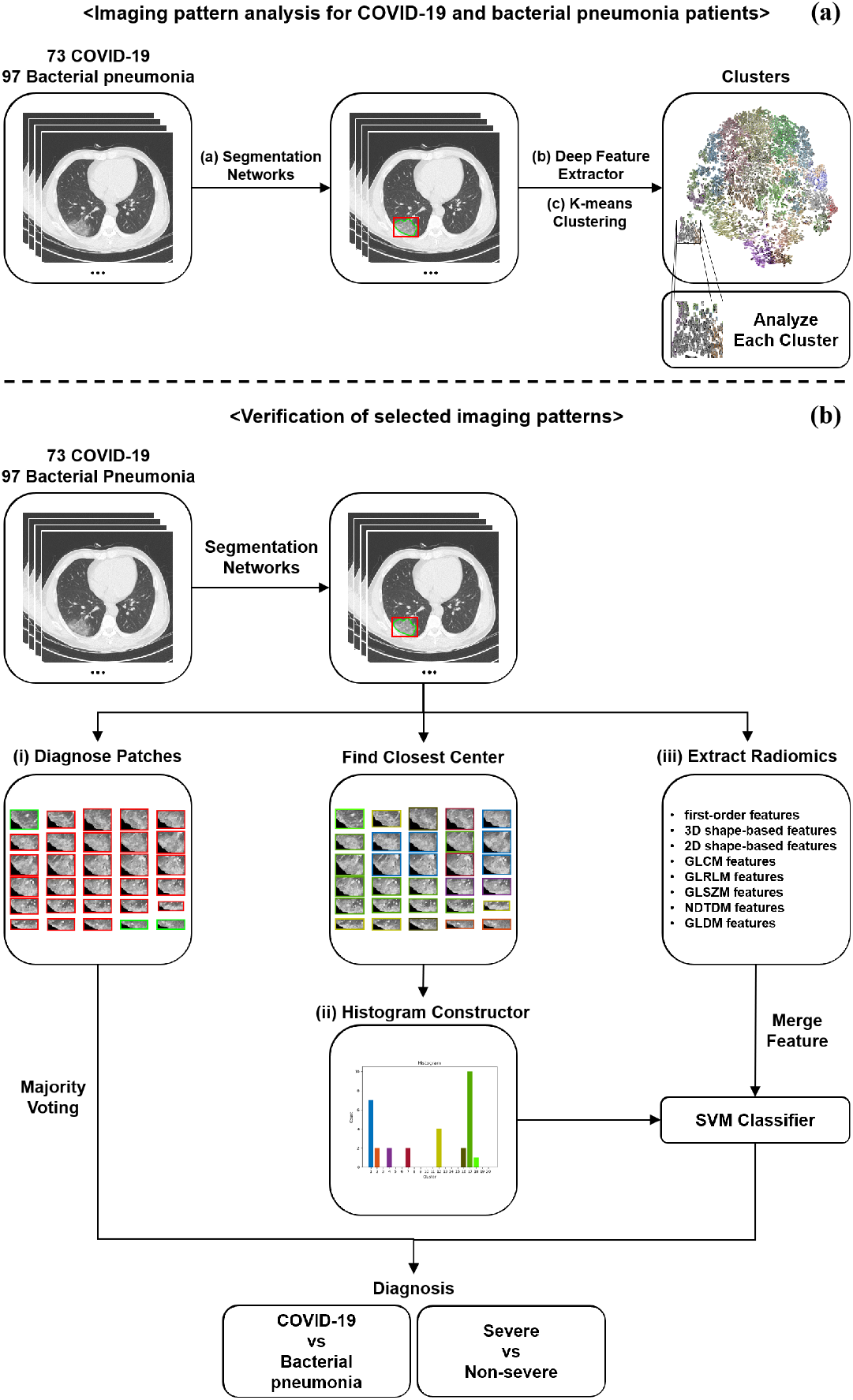
Diagram of the modeling framework. For analyzing the imaging patterns, the segmentation models were employed to extract regions of interest of the lung and lesions in the chest CT scans. Then, the ResNet50 model takes lesion cropped patches as input and returns a label prediction and a 2048-dimensional feature vector. Next, the K-means clustering algorithm was used to cluster 2D lesion features into 20 groups. To verify the usability of these patterns, (i) majority voting; (ii) constructed histogram with SVM; and (iii) Radiomics features (with/without constructed histogram) with SVM were employed. Disease and severity classification tasks were performed.

A Mask-cascade-RCNN-ResNeSt-200 with deformable convolution neural network (DCN) architecture was employed in module (a) to extract the lung and lesion regions in the chest CT scans [14], [15]. The lung segmentation model was trained and evaluated on a total 51,978 manually segmented slices (train: 50,756, test: 1,222) from two public datasets (NSCLC, 20cases) [16], [17]. For the lesion segmentation task, 6,971 manually segmented slices (train: 5,854, test: 1,117) from three publicly available datasets were used i.e. (20cases, MosMed, MSD) [17]–[19]. Then, for the data from Yeungnam University Medical Center, lesions were extracted using the trained models. The patches smaller than 13mm were not used to avoid misclassification caused by wrong segmentation or noise such as motion artifacts, as this may have a negative effect on subsequent analysis.

The deep neural network in module (b) employed a ResNet50 model trained to differentiate lesion patches of COVID-19 from those of bacterial pneumonia patients [20]. The model took a lesion cropped patch as input and returned a label prediction. A total of 12,235 lesion patches (train: 6,181, test: 6,054) from 170 patients were employed. A 2048-dimensional feature vector extracted from the intermediate layer of the ResNet50 model used in the clustering phase, i.e., module (c).

In module (c), the K-means algorithm was applied to cluster the lesion features into 20 groups. To profile the typical or relatively atypical imaging features of COVID-19, a total of 12,235 lesion patches from 170 patients were represented in two-dimensional space via a t-distributed Stochastic Neighbor Embedding based reduction of the 2048-dimensional feature vectors [21]. The lesion images in each cluster initially grouped by K-means were later manually described by one radiologist (Jongsoo Park) using imaging terms, wherein three pulmonologists (Kyung Soo Hong, Jong Geol Jang, and June Hong Ahn) evaluated the descriptions and reached a consensus.

### C. VERIFICATION OF SELECTED IMAGING PATTERNS

Patient-level diagnosis can be achieved by aggregating the lesion-level predictions using majority voting. However, in this way, it is difficult to analyze what patterns the patient has and which combination of patterns are highly correlated with the disease or severity. To further quantify the diagnostic performance of the extracted imaging patterns, we constructed a histogram of lesion types using the clusters obtained via module (c) for each CT. Figure 2(b) shows a diagram of the evaluation framework. In addition, the constructed histograms were analyzed using an independent two-sample t-test to evaluate the significance of each cluster regarding: (i) COVID-19 and bacterial pneumonia patients, (ii) severe and non-severe cases. A two-tailed P <0.05 was taken to indicate statistical significance. All statistical analyses were performed using Scipy (1.5.0, https://www.scipy.org/). Furthermore, we compared the mean histograms to confirm cluster relevance using the mean of each histogram in each group i.e. COVID-19, bacterial pneumonia, severe, and nonsevere. The cluster which showed high statistical significance (P <0.001) indicated a significant value differences when performing mean histogram comparison.

Furthermore, we compared the classification accuracy between majority voting of the lesion-level predictions and SVM based inference which was trained on a combination of the proposed histogram with radiomics features. A total of 107 features were extracted using radiomics from each CT volume, including first-order statistics, shape-based features, etc. Finally, 20 features from our proposed histogram and 107 features from radiomics features were combined. We compared the performance with/without the combination of the radiomic features to verify effectiveness.

## III. RESULTS

### A. DEMOGRAPHIC AND CLINICAL CHARACTERISTICS

Hospitalized patients with confirmed COVID-19 (73 patients) and bacterial pneumonia (97 patients) were included in this study (Fig. 1). Baseline characteristics of all patients are summarized in Table 1. The patients in the COVID-19 group were older than the patients in the bacterial pneumonia group (58.70 ± 16.49 vs. 39.86 ± 8.69, p <0.001). 36 patients (49.3%) of COVID-19 and 55 patients (56.7%) of bacterial pneumonia were male. Body temperature (37.21 ± 0.67 vs. 37.76 ± 0.92, p <0.001), and heart rate (86.70 ± 13.94 vs. 94.37 ± 16.91, p =0.020) were significantly lower in patients with COVID-19. Systolic blood pressure (128.55 ± 19.44 vs. 115.71 ± 19.42, p <0.001), and diastolic blood pressure (80.74 ± 12.15 vs. 71.42 ± 13.52, p <0.001) were significantly higher for COVID-19 patients.

**TABLE 1.** Characteristics of the study participants with COVID-19 and bacterial pneumonia.

| Variables | COVID-19 (n=73) | Bacterial pneumonia (n=97) | P-value |
| --- | --- | --- | --- |
| Age, yr | 58.70 (16.49) | 39.86 (8.69) | 0.0000 |
| Male, sex | 36 (49.3) | 55 (56.7) | 0.3903 |
| Vital signs on admission |  |  |  |
| Body temperature, °C | 37.21 (0.67) | 37.76 (0.92) | 0.0000 |
| Heart rate, beats/min | 86.70 (13.94) | 94.37 (16.91) | 0.0020 |
| Systolic BP, mm Hg | 128.55 (19.44) | 115.71 (19.42) | 0.0000 |
| Diastolic BP, mm Hg | 80.74 (12.15) | 71.42 (13.52) | 0.0000 |

Data are presented as the mean ± standard deviation or number (%)

### B. DEEP CHEST CT MANIFESTATIONS

We summarize the deep chest CT manifestations observed in Table 2, with lesion patches visualized in Figure 3, respectively. Typical COVID-19 CT manifestations such as GGOs with interlobular septal thickening in the peripheral lungs were observed in clusters 4 and 17 (See Figure 4(a)). On the other hand, manifestations typical to bacterial pneumonia such as crazy-paving appearance in the posterior lungs were observed in clusters 5 and 10, as presented in Figure 4(b). These typical clusters showed P-values less than 0.001.

**TABLE 2.**
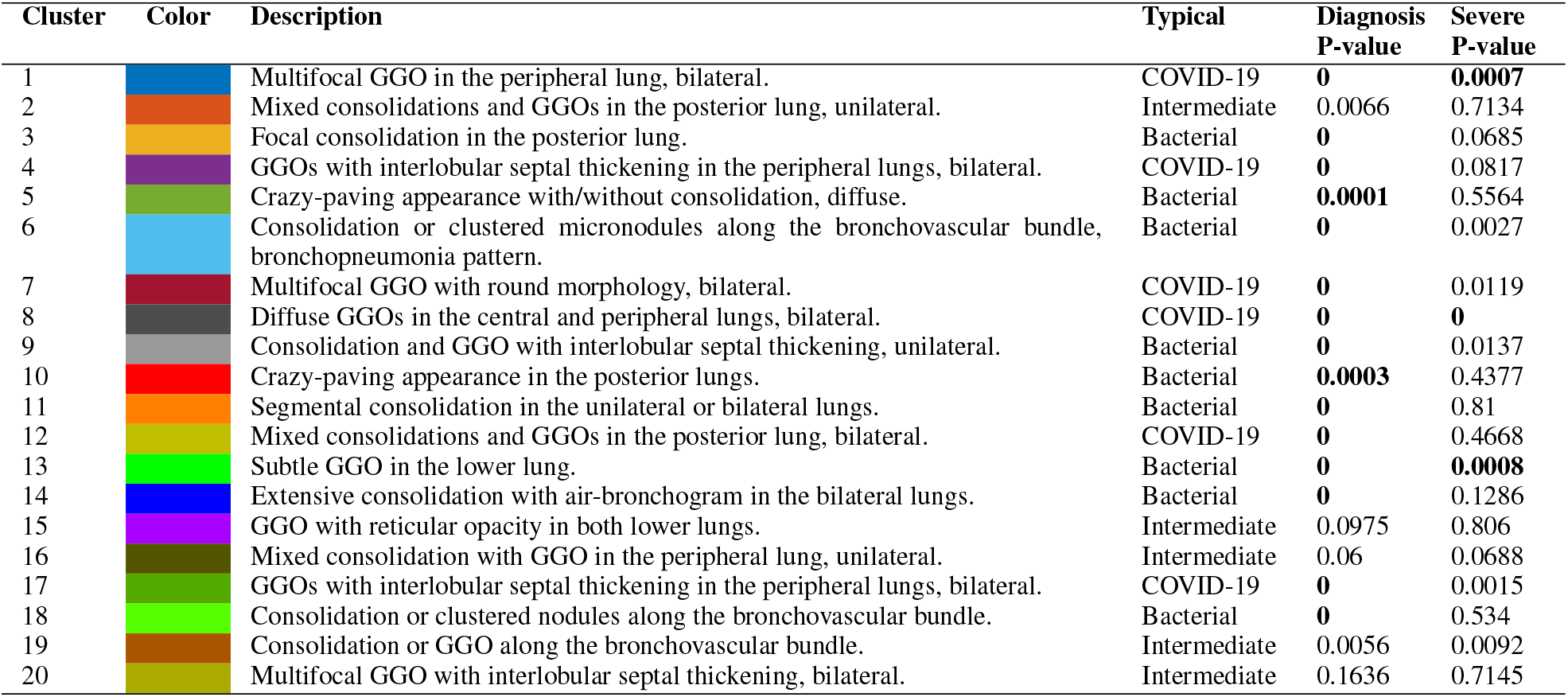
Summary of lesion cluster with imaging description term and P-value results. The t-test were performed for diseased and severe groups, respectively.

| Cluster | Color | Description | Typical | Diagnosis P-value | Severe P-value |
| --- | --- | --- | --- | --- | --- |
| 1 |  | Multifocal GGO in the peripheral lung, bilateral. | COVID-19 | **0** | **0.0007** |
| 2 |  | Mixed consolidations and GGOs in the posterior lung, unilateral. | Intermediate | 0.0066 | 0.7134 |
| 3 |  | Focal consolidation in the posterior lung. | Bacterial | **0** | 0.0685 |
| 4 |  | GGOs with interlobular septal thickening in the peripheral lungs, bilateral. | COVID-19 | **0** | 0.0817 |
| 5 |  | Crazy-paving appearance with/without consolidation, diffuse. | Bacterial | **0.0001** | 0.5564 |
| 6 |  | Consolidation or clustered micronodules along the bronchovascular bundle, bronchopneumonia pattern. | Bacterial | **0** | 0.0027 |
| 7 |  | Multifocal GGO with round morphology, bilateral. | COVID-19 | **0** | 0.0119 |
| 8 |  | Diffuse GGOs in the central and peripheral lungs, bilateral. | COVID-19 | **0** | **0** |
| 9 |  | Consolidation and GGO with interlobular septal thickening, unilateral. | Bacterial | **0** | 0.0137 |
| 10 |  | Crazy-paving appearance in the posterior lungs. | Bacterial | **0.0003** | 0.4377 |
| 11 |  | Segmental consolidation in the unilateral or bilateral lungs. | Bacterial | **0** | 0.81 |
| 12 |  | Mixed consolidations and GGOs in the posterior lung, bilateral. | COVID-19 | **0** | 0.4668 |
| 13 |  | Subtle GGO in the lower lung. | Bacterial | **0** | **0.0008** |
| 14 |  | Extensive consolidation with air-bronchogram in the bilateral lungs. | Bacterial | **0** | 0.1286 |
| 15 |  | GGO with reticular opacity in both lower lungs. | Intermediate | 0.0975 | 0.806 |
| 16 |  | Mixed consolidation with GGO in the peripheral lung, unilateral. | Intermediate | 0.06 | 0.0688 |
| 17 |  | GGOs with interlobular septal thickening in the peripheral lungs, bilateral. | COVID-19 | **0** | 0.0015 |
| 18 |  | Consolidation or clustered nodules along the bronchovascular bundle. | Bacterial | **0** | 0.534 |
| 19 |  | Consolidation or GGO along the bronchovascular bundle. | Intermediate | 0.0056 | 0.0092 |
| 20 |  | Multifocal GGO with interlobular septal thickening, bilateral. | Intermediate | 0.1636 | 0.7145 |

**FIGURE 3.**
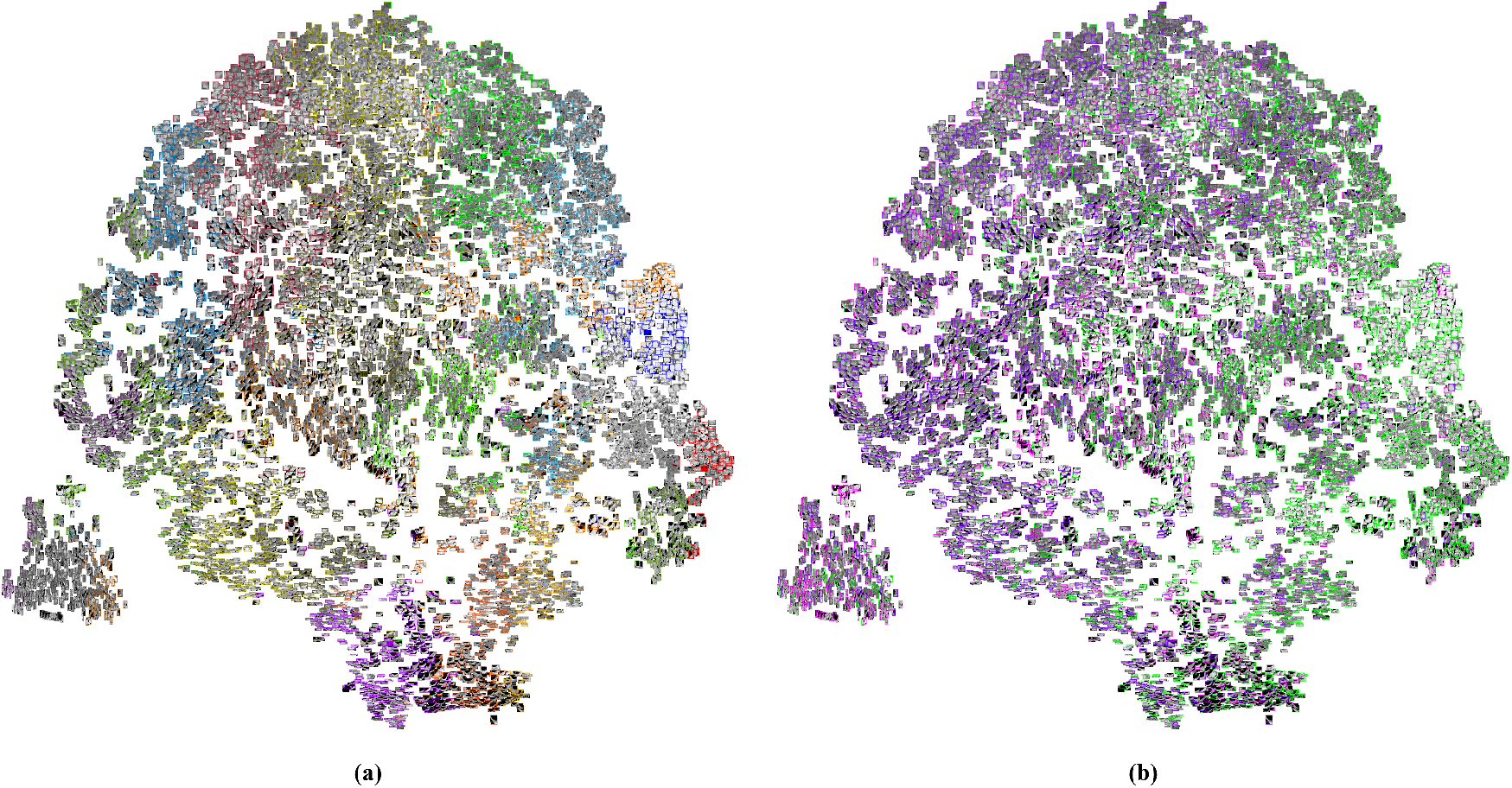
Visualization of the lesion patches from 170 patients into two-dimensions. (a) K-means clusters (20 groups) were used for distinction. Different color edge represents different groups. (b) True diagnosis was used for distinction. Purple color represents non-severe COVID-19, magenta represents severe COVID-19, and green for bacterial pneumonia.

**FIGURE 4.**
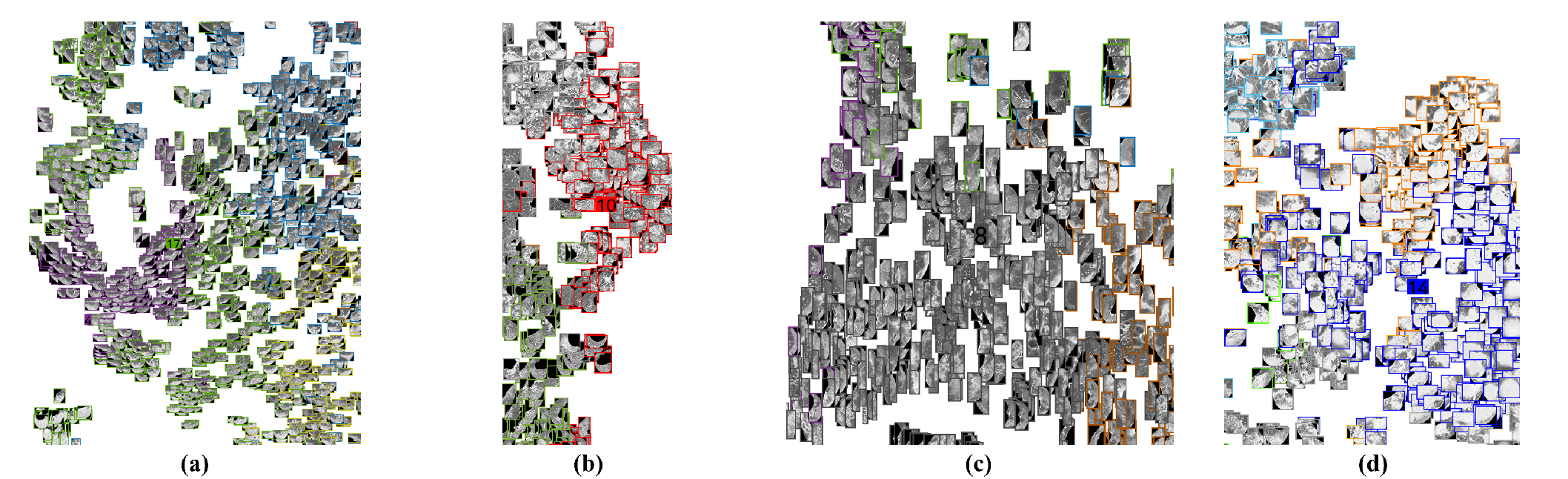
(a) Typical COVID-19 CT manifestations - GGOs with interlobular septal thickening in the peripheral lungs - were observed in cluster 4 and 17. (b) bacterial pneumonia CT manifestation – crazy-paving appearance in the posterior lungs – observed in cluster 5 and 10. (c) Cluster 8 showed the lowest t-test P-value <0.001 for diseased and severe groups which is the typical pattern for severe COVID-19 patients i.e. diffuse GGOs in the central and peripheral lungs. (d) Bacterial pneumonia CT manifestation – extensive consolidation with air-bronchogram – was observed in cluster 14. These clusters showed a t-test P-value <0.001. Lesions were colored based on K-means clustering result.

Notably, 15 out of the 20 clusters showed key characteristics for the discrimination of COVID-19 from bacterial pneumonia with P-values less than 0.001. Among 15 clusters, three clusters with P-values less than 0.001 were shown to be key for severity classification. Moreover, two clusters could classify both COVID-19 and severe patients, with one of the clusters (#8) showed diffuse GGOs in the central and peripheral lungs which represents a typical severe COVID-19 CT manifestation. Figure 4(c) presents the severe COVID-19 patient’s CT manifestations.

In principle, the observed relative differences between lesion clusters are key to better understand the entire lesion distribution. As shown in Figure 4(d), extensive consolidation with air-bronchogram was observed in severe bacterial pneumonia patients. Compared with the severe COVID-19 patients who mainly showed diffuse GGOs, we could easily distinguish the difference in patterns observed in the later stages of disease onset.

To confirm cluster relevance, we compared the mean histograms of different tasks as presented in Figure 5. The clusters showed high significance (P <0.001) in Table 2 also showed a significantly different values in mean histogram comparison. Notably, the mean histogram of COVID-19 can be discriminated from bacterial pneumonia by comparing the considerably increased values in cluster 1, 4, 7, 12, and 17. On the other hand, the mean histogram of bacterial pneumonia can be discriminated from COVID-19 by comparing the significantly increased values of cluster 3, 6, 9, 11, 13, 14, and 18. In addition, the mean histogram of the severe COVID-19 patient shows consistently increased values in cluster 8 compared to the mean histogram of the non-severe COVID-19 patient. This indicates that the diagnosis is highly relevant to the presence of a lesion in a certain cluster, and the patient with a lesion in cluster 8 can be considered as a more severe COVID-19 patient than patients with lesions in other clusters. In summary, the cluster interpreted by the radiologist as being highly relevant to typical COVID-19 and bacterial pneumonia patterns showed high correlation with P-value significance in statistical analyses, with notable differences observed in mean histogram comparison.

**FIGURE 5.**
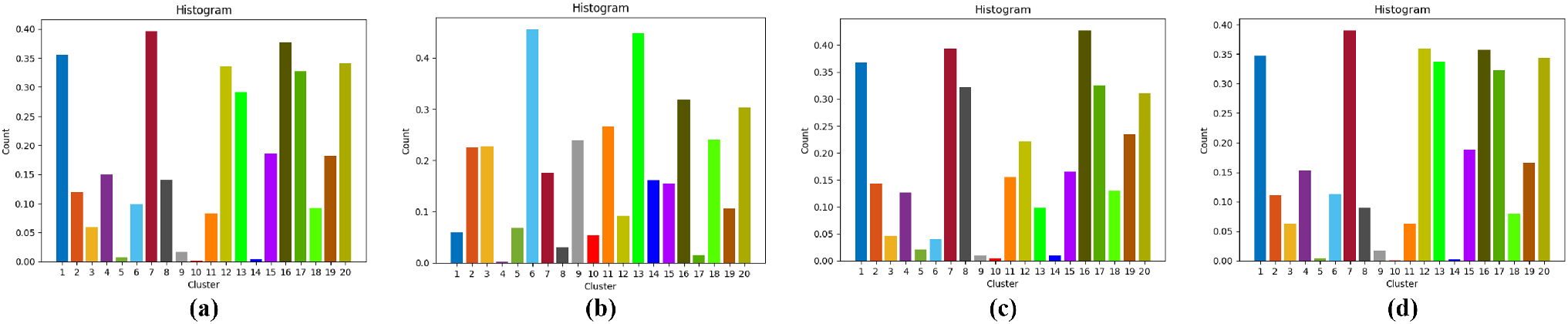
Mean histograms i.e. (a) mean histogram of COVID-19; (b) mean histogram of bacterial pneumonia; (c) mean histogram of severe COVID-19 patients; (d) mean histogram of non-severe COVID-19 patients.

### C. DISEASE AND SEVERITY CLASSIFICATION

In Table 3, we present disease classification accuracy across several metrics under different settings. First, the baseline method based on majority voting of predictions achieved 87.5% accuracy for COVID-19 patient classification. For the SVM classifier, we consider two scenarios, (i) with histogram features only or (ii) with radiomic features only. Notably, the classifier trained under the first setting achieved 88.7% accuracy; a minor improvement over the baseline, whereas in setting (ii) - 81.25% was reported, a considerable decrease from the other models. However, when the SVM classifier was trained with both radiomics and histogram features, accuracy significantly improved to 91.2% from 87.5%. This further shows that the features learned by deep learning are highly robust for accurate patient diagnosis. In addition, the constructed histograms can accurately express the correlation of lesion features observed in each patient and/or can represent some combination of patterns that could lead to a severe outcome of a disease diagnosis. In comparison to the naïve aggregation method baseline i.e. majority voting, often highly limited in patient-level representations; the proposed deep feature representation highlights several key advantages.

**TABLE 3.** Test accuracy of COVID-19 diagnosis.

| Model | Accuracy (%) | Sensitivity (%) | Specificity (%) |
| --- | --- | --- | --- |
| Majority Voting | 87.5 (70 of 80) | 92.5 (37 of 40) | 82.5 (33 of 40) |
| Radiomics only | 81.25 (65 of 80) | 87.5 (35 of 40) | 75 (30 of 40) |
| Histogram only | 88.7 (71 of 80) | 85 (34 of 40) | 92.5 (37 of 40) |
| Histogram with Radiomics | **91.2 (73 of 80)** | 85 (34 of 40) | 97.5 (39 of 40) |

Table 4 shows the severity classification accuracy. In terms of accuracy alone, SVM classifiers that only use either histogram or radiomics only achieved the same performance, i.e. 82.5% for both, with key differences noted in the sensitivity and specificity of the models. On the other hand, when the features were combined considerable improvement (+12.5%) was noted i.e. 95% accuracy, surpassing previous models. This may be attributed to the fact that the histogram is not able to represent the absolute size difference of lesions, thus the combination mitigates the issue. Rather than using a single feature alone, the combination of features proved to be invaluable for diagnosis.

**TABLE 4.** Test accuracy of severity COVID-19 classification.

| Model | Accuracy (%) | Sensitivity (%) | Specificity (%) |
| --- | --- | --- | --- |
| Radiomics only | 82.5 (33 of 40) | 66.67 (4 of 6) | 85.29 (28 of 34) |
| Histogram only | 82.5 (33 of 40) | 50 (3 of 6) | 88.24 (30 of 34) |
| Histogram with Radiomics | **95 (38 of 40)** | 83.3 (5 of 6) | 97.06 (33 of 34) |

### D. LUNG AND LESION SEGMENTATION

The lung and lesion segmentation model achieved a dice coefficient score of 97.18%, and 78.06%, respectively. Figure 6 shows the lesion segmentation results around the average dice coefficient score of 78.06%, which helps to qualitatively understand the accuracy of lesion segmentation.

**FIGURE 6.**
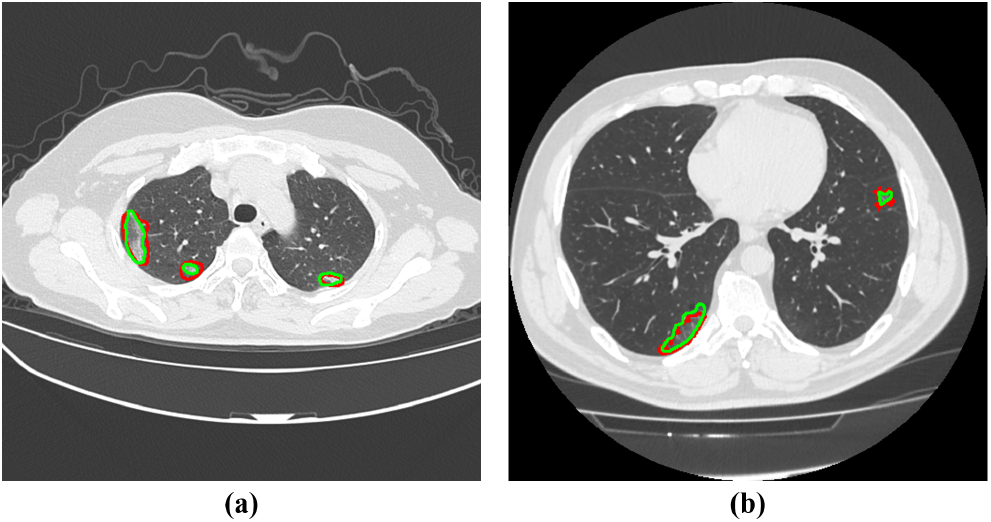
Representative slices which showed an average dice coefficient score (78.06%) for lesion segmentation i.e. (a) slice showed a dice coefficient score of 77.08%; (b) slice showed a dice coefficient score of 78.31%. Red color represents a manual segmentation by human and green color represents an automated segmentation by the trained segmentation model. Due to the ambiguity of lesion boundaries, an average dice score of 78.06% showed satisfactory lesion segmentation results for the deep feature extractor.

## IV. DISCUSSION

Our deep chest CT analysis is in accordance with various CT findings of COVID-19 patients reported in literature. Chung et al. reported that typical CT findings of COVID-19 include bilateral pulmonary GGO and consolidative opacities which sometimes have a rounded morphology and are distributed in the peripheral lung [22]. Song et al. reported that pure GGO or GGO with reticular and/or interlobular septal thickening with predominant distribution in the posterior or peripheral lung involvements were observed in COVID-19 patients [5]. Caruso et al. highlighted the presence of the peripheral GGOs associated with multilobe and posterior lung involvement in COVID-19 patients in Italy [23]. Similar patterns were also observed in our research i.e. GGO distributed in the peripheral lungs in cluster 4 and 17, with multifocal GGOs with round morphology in cluster 1 and 7, respectively. Regarding the common findings of severe COVID-19, Pan et al. report that an increase in GGO, consolidative opacities, and interstitial septal thickening was noted, and Song et al. reported that a significantly more GGOs including pure GGO and a GGO with reticular and/or interlobular septal thickening was observed in the later stages of COVID-19 [5], [24]. An increase in GGOs in the central and peripheral lung was observed in our study in cluster 8, therefore, the published CT findings of severe COVID-19 patients show high consistency with our study.

Radiomic features are considered a useful general purpose analysis technique, i.e. for distinguishing the lung nodules (malignant versus benign) or hospital stay (severity) prediction [25], [26]. However, several limitations exist and features alone are often insufficient to distinguish between diseases when subtle radiological differences are observed in the image. Discriminating COVID-19 from bacterial pneumonia is regarded as one of the exemplars of such challenges. Here, our method shows the benefit of using deep learning to obtain more robust representations that are more clinically relevant to key imaging characteristics for COVID-19 diagnosis. We quantitatively show that the constructed histogram better captures the overall statistics of the lesion features. Moreover, the SVM classifier can diagnose diseases or patient’s severity more accurately than the radiomics features alone.

Our method has two notable advantages compared to common deep learning algorithms; interpretability and generalized representation. Common deep learning methods are limited in interpretability even though they can visualize the important regions using heatmaps [27], [28]. Our method can explain the reasons of diagnosis by checking the presence of specific patterns represented in the patient’s histogram. We verified the key patterns with mean histograms for each disease and severity group and found the important key diagnostic imaging patterns in accordance with published literature. This indicates that our method is safer and more transparent for medical assistance. Moreover, although the proposed feature learning model was not trained to classify severity among patients, the obtained features are fairly generalized for severity classification. The imaging features were divided into 20 clusters and verified by radiologists using imaging terms i.e. an independent representation of a specific diagnosis. The constructed histogram can be used for general diagnosis regardless of the trained diseases, thus the constructed histogram showed considerable diagnostic accuracy for severity classification.

The proposed framework was trained only on a single institute cohort, therefore current models may fail to accurately represent unobserved cohort CT manifestations. Moreover, the demographic characteristics of the disease group showed statistical significance. Even though these differences do not affect the CT imaging features, our method will be evaluated on multi center data in future for verification. Additionally, the K-means algorithm requires an arbitrary number of groups, it was set to 20 in this study even though it may not be the optimal value. Applying better clustering algorithms and selecting an optimal number of clusters will be the subject of future research.

## V. CONCLUSION

In conclusion, the CT images of COVID-19 in Daegu, Korea were grouped into 20 clusters. These groups were analyzed and compared with the patterns described in literature. To verify the effectiveness of these clusters, we performed two classification tasks by constructing histograms from the clusters. We confirmed the correlations of the image patterns extracted by the proposed method are more relevant to the clinical setting than the common methods which use radiomics or naïve deep features.

## Data Availability

N/A

## APPENDIX A

**TABLE 5.**
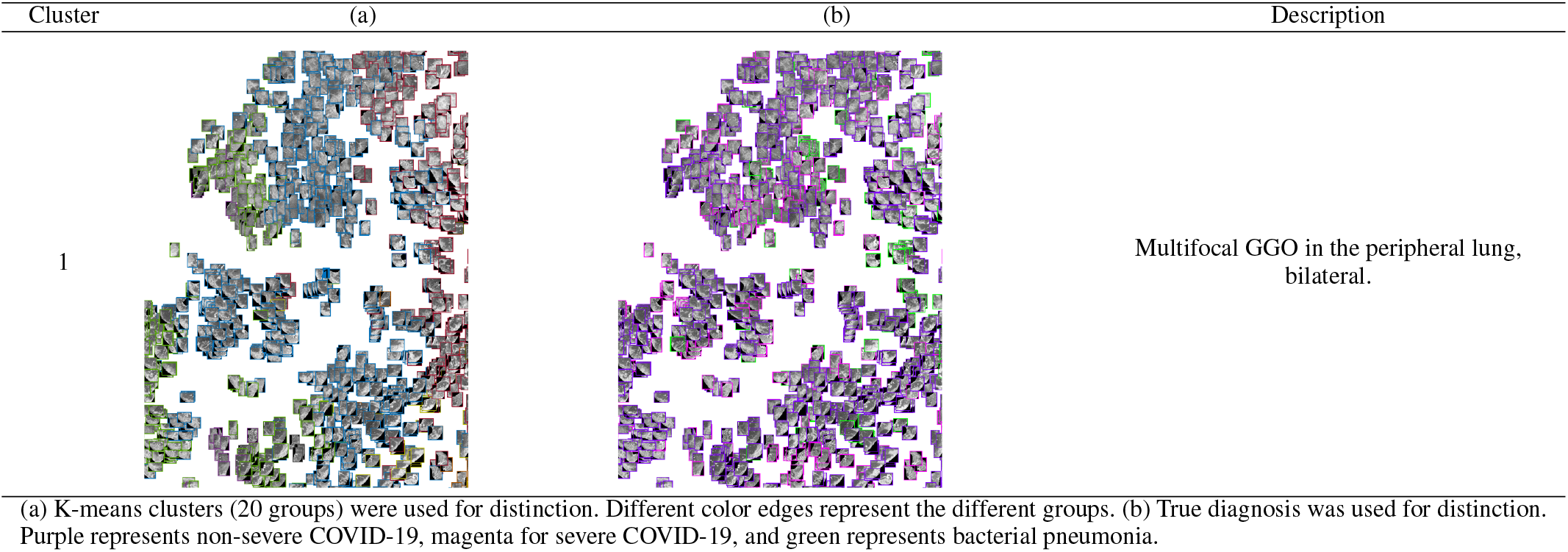
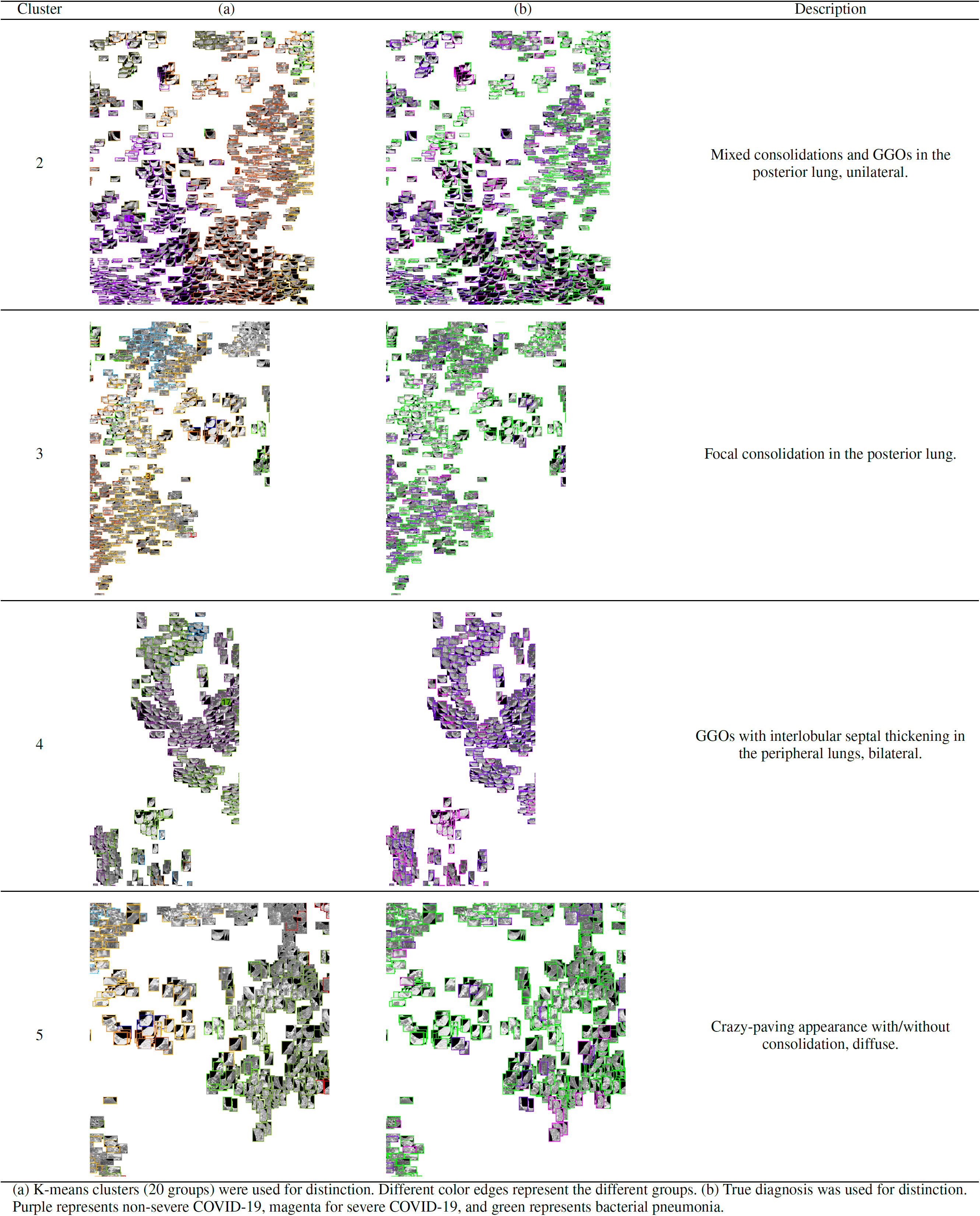
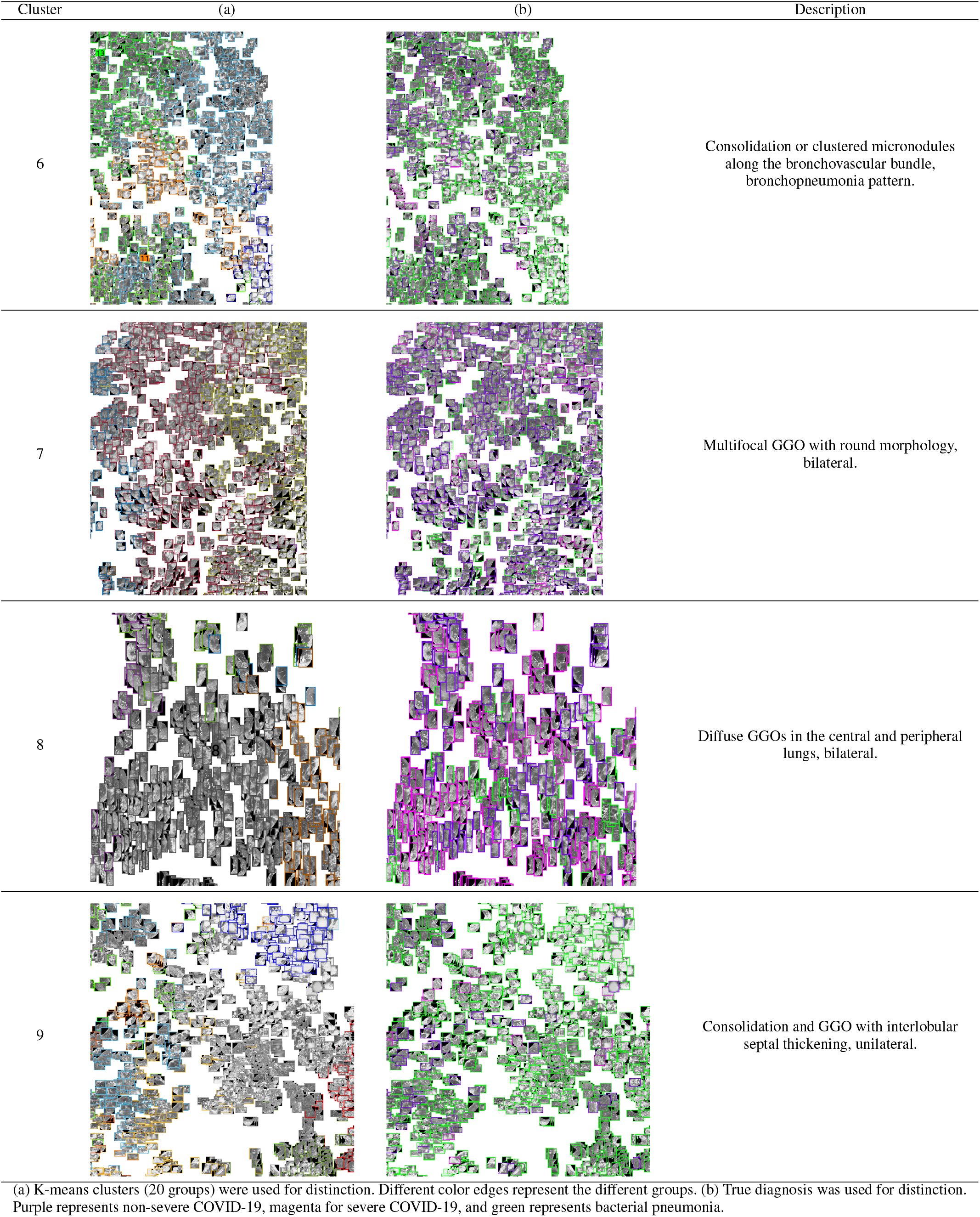
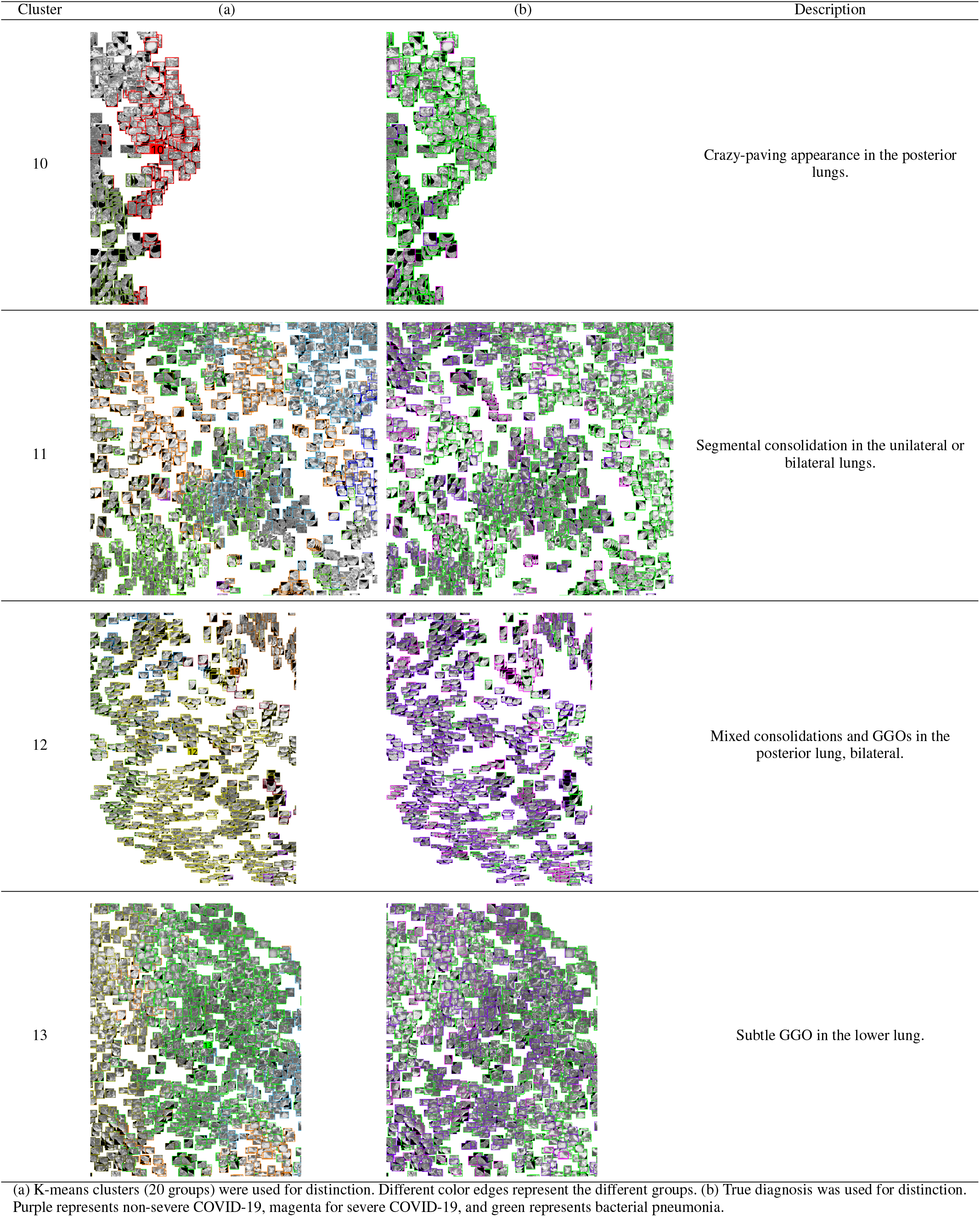
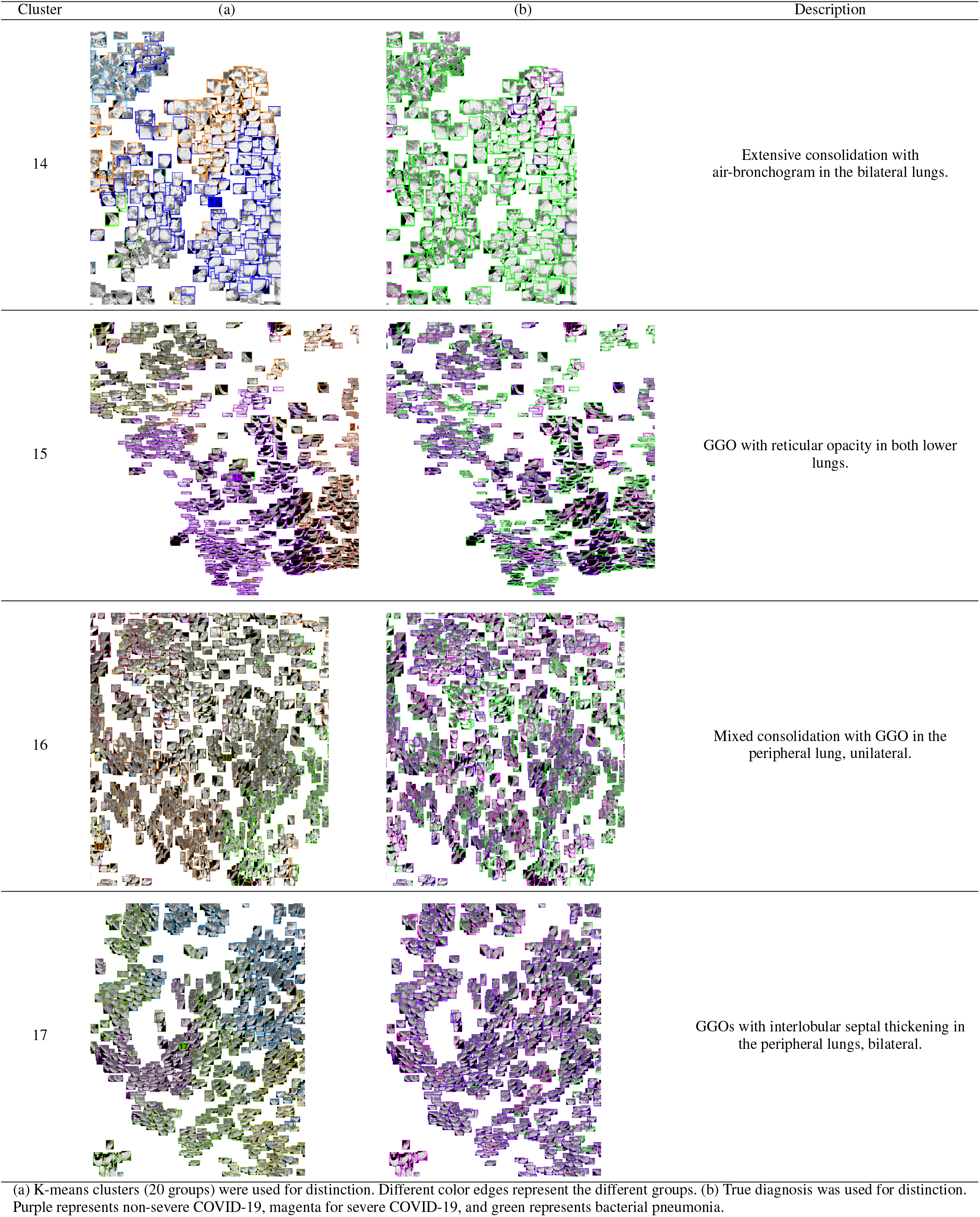
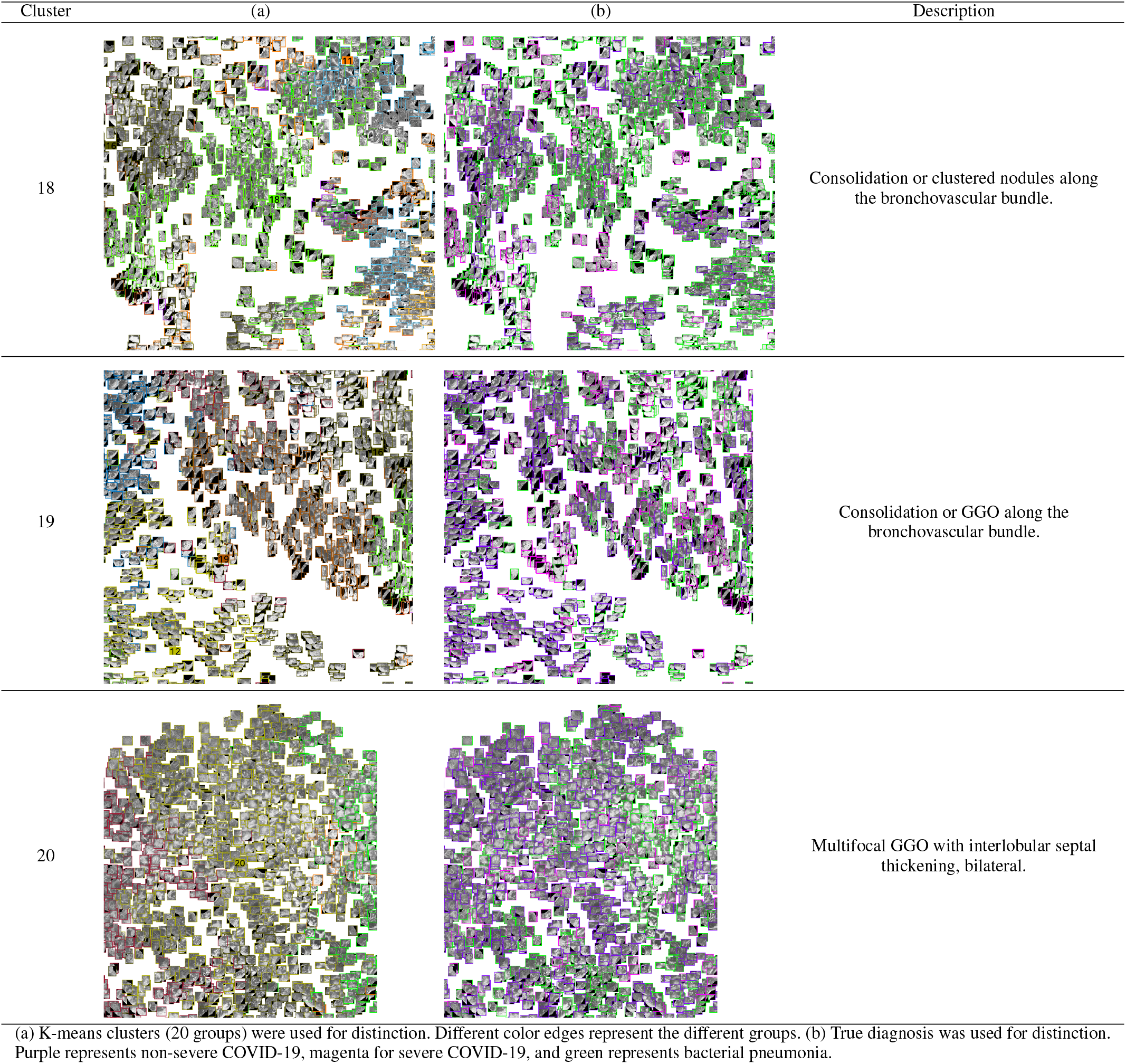
Zoomed visualization of the lesion patches.

| Cluster | (a) | (b) | Description |
| --- | --- | --- | --- |
| 1 |  |  | Multifocal GGO in the peripheral lung, bilateral. |
| (a) K-means clusters (20 groups) were used for distinction. Different color edges represent the different groups. (b) True diagnosis was used for distinction. Purple represents non-severe COVID-19, magenta for severe COVID-19, and green represents bacterial pneumonia. |  |  |  |
| 2 |  |  | Mixed consolidations and GGOs in the posterior lung, unilateral. |
| 3 |  |  | Focal consolidation in the posterior lung. |
| 4 |  |  | GGOs with interlobular septal thickening in the peripheral lungs, bilateral. |
| 5 |  |  | Crazy-paving appearance with/without consolidation, diffuse. |
| 6 |  |  | Consolidation or clustered micronodules along the bronchovascular bundle, bronchopneumonia pattern. |
| 7 |  |  | Multifocal GGO with round morphology, bilateral. |
| 8 |  |  | Diffuse GGOs in the central and peripheral lungs, bilateral. |
| 9 |  |  | Consolidation and GGO with interlobular septal thickening, unilateral. |
| 10 |  |  | Crazy-paving appearance in the posterior lungs. |
| 11 |  |  | Segmental consolidation in the unilateral or bilateral lungs. |
| 12 |  |  | Mixed consolidations and GGOs in the posterior lung, bilateral. |
| 13 |  |  | Subtle GGO in the lower lung. |
| 14 |  |  | Extensive consolidation with air-bronchogram in the bilateral lungs. |
| 15 |  |  | GGO with reticular opacity in both lower lungs. |
| 16 |  |  | Mixed consolidation with GGO in the peripheral lung, unilateral. |
| 17 |  |  | GGOs with interlobular septal thickening in the peripheral lungs, bilateral. |
| 18 |  |  | Consolidation or clustered nodules along the bronchovascular bundle. |
| 19 |  |  | Consolidation or GGO along the bronchovascular bundle. |
| 20 |  |  | Multifocal GGO with interlobular septal thickening, bilateral. |

(a) K-means clusters (20 groups) were used for distinction. Different color edges represent the different groups. (b) True diagnosis was used for distinction. Purple represents non-severe COVID-19, magenta for severe COVID-19, and green represents bacterial pneumonia.

## REFERENCES

[1] W.-j. Guan, Z.-y. Ni, Y. Hu, W.-h. Liang, C.-q. Ou, J.-x. He, L. Liu, H. Shan, C.-l. Lei, D. S. Hui et al., “Clinical characteristics of coronavirus disease 2019 in china,” New England journal of medicine, vol. 382, no. 18, pp. 1708–1720, 2020.

[2] X. Xie, Z. Zhong, W. Zhao, C. Zheng, F. Wang, and J. Liu, “Chest ct for typical 2019-ncov pneumonia: relationship to negative rt-pcr testing,” Radiology, p. 200343, 2020.

[3] P. Huang, T. Liu, L. Huang, H. Liu, M. Lei, W. Xu, X. Hu, J. Chen, and B. Liu, “Use of chest ct in combination with negative rt-pcr assay for the 2019 novel coronavirus but high clinical suspicion,” Radiology, vol. 295, no. 1, pp. 22–23, 2020.

[4] Y. Fang, H. Zhang, J. Xie, M. Lin, L. Ying, P. Pang, and W. Ji, “Sensitivity of chest ct for covid-19: comparison to rt-pcr,” Radiology, p. 200432, 2020.

[5] F. Song, N. Shi, F. Shan, Z. Zhang, J. Shen, H. Lu, Y. Ling, Y. Jiang, and Y. Shi, “Emerging 2019 novel coronavirus (2019-ncov) pneumonia,” Radiology, vol. 295, no. 1, pp. 210–217, 2020.

[6] S. Salehi, A. Abedi, S. Balakrishnan, and A. Gholamrezanezhad, “Coronavirus disease 2019 (covid-19): a systematic review of imaging findings in 919 patients,” American Journal of Roentgenology, pp. 1–7, 2020.

[7] S. Zhou, Y. Wang, T. Zhu, and L. Xia, “Ct features of coronavirus disease 2019 (covid-19) pneumonia in 62 patients in wuhan, china,” American Journal of Roentgenology, vol. 214, no. 6, pp. 1287–1294, 2020.

[8] H. X. Bai, B. Hsieh, Z. Xiong, K. Halsey, J. W. Choi, T. M. L. Tran Pan, L.-B. Shi, D.-C. Wang, J. Mei et al., “Performance of radiologists in differentiating covid-19 from viral pneumonia on chest ct,” Radiology, p. 200823, 2020.

[9] H. Choi, X. Qi, S. H. Yoon, S. J. Park, K. H. Lee, J. Y. Kim, Y. K. Lee, H. Ko, K. H. Kim, C. M. Park et al., “Extension of coronavirus disease 2019 (covid-19) on chest ct and implications for chest radiograph interpretation,” Radiology: Cardiothoracic Imaging, vol. 2, no. 2, p. e200107, 2020.

[10] D. M. Hansell, A. A. Bankier, H. MacMahon, T. C. McLoud, N. L. Muller, and J. Remy, “Fleischner society: glossary of terms for thoracic imaging,” Radiology, vol. 246, no. 3, pp. 697–722, 2008.

[11] G. Litjens, T. Kooi, B. E. Bejnordi, A. A. A. Setio, F. Ciompi, M. Ghafoorian, J. A. Van Der Laak, B. Van Ginneken, and C. I. Sánchez, “A survey on deep learning in medical image analysis,” Medical image analysis, vol. 42, pp. 60–88, 2017.

[12] S. Lloyd, “Least squares quantization in pcm,” IEEE transactions on information theory, vol. 28, no. 2, pp. 129–137, 1982.

[13] A. D. T. Force, V. Ranieri, G. Rubenfeld, B. Thompson, N. Ferguson, E. Caldwell et al., “Acute respiratory distress syndrome,” Jama, vol. 307, no. 23, pp. 2526–2533, 2012.

[14] H. Zhang, C. Wu, Z. Zhang, Y. Zhu, Z. Zhang, H. Lin, Y. Sun, T. He, J. Mueller, R. Manmatha et al., “Resnest: Split-attention networks,” arXiv preprint arXiv:2004.08955, 2020.

[15] J. Dai, H. Qi, Y. Xiong, Y. Li, G. Zhang, H. Hu, and Y. Wei, “Deformable convolutional networks,” in Proceedings of the IEEE international conference on computer vision, 2017, pp. 764–773.

[16] H. J. Aerts, E. R. Velazquez, R. T. Leijenaar, C. Parmar, P. Grossmann, S. Carvalho, J. Bussink, R. Monshouwer, B. Haibe-Kains, D. Rietveld et al., “Decoding tumour phenotype by noninvasive imaging using a quantitative radiomics approach,” Nature communications, vol. 5, no. 1, pp. 1–9, 2014.

[17] M. Jun, G. Cheng, W. Yixin, A. Xingle, G. Jiantao, Y. Ziqi, Z. Minqing, L. Xin, D. Xueyuan, C. Shucheng et al., “Covid-19 ct lung and infection segmentation dataset,” Zenodo, Apr, vol. 20, 2020.

[18] S. Morozov, A. Andreychenko, N. Pavlov, A. Vladzymyrskyy, N. Ledikhova, V. Gombolevskiy, I. A. Blokhin, P. Gelezhe, A. Gonchar, and V. Y. Chernina, “Mosmeddata: Chest ct scans with covid-19 related findings dataset,” arXiv preprint arXiv:2005.06465, 2020.

[19] A. L. Simpson, M. Antonelli, S. Bakas, M. Bilello, K. Farahani, B. Van Ginneken, A. Kopp-Schneider, B. A. Landman, G. Litjens, B. Menze et al., “A large annotated medical image dataset for the development and evaluation of segmentation algorithms,” arXiv preprint arXiv:1902.09063, 2019.

[20] K. He, X. Zhang, S. Ren, and J. Sun, “Deep residual learning for image recognition,” in Proceedings of the IEEE conference on computer vision and pattern recognition, 2016, pp. 770–778.

[21] L. v. d. Maaten and G. Hinton, “Visualizing data using t-sne,” Journal of machine learning research, vol. 9, no. Nov, pp. 2579–2605, 2008.

[22] M. Chung, A. Bernheim, X. Mei, N. Zhang, M. Huang, X. Zeng, J. Cui, W. Xu, Y. Yang, Z. A. Fayad et al., “Ct imaging features of 2019 novel coronavirus (2019-ncov),” Radiology, vol. 295, no. 1, pp. 202–207, 2020.

[23] D. Caruso, M. Zerunian, M. Polici, F. Pucciarelli, T. Polidori, C. Rucci, G. Guido, B. Bracci, C. de Dominicis, and A. Laghi, “Chest ct features of covid-19 in rome, italy,” Radiology, p. 201237, 2020.

[24] Y. Pan, H. Guan, S. Zhou, Y. Wang, Q. Li, T. Zhu, Q. Hu, and L. Xia, “Initial ct findings and temporal changes in patients with the novel coronavirus pneumonia (2019-ncov): a study of 63 patients in wuhan, china,” European radiology, pp. 1–4, 2020.

[25] X. Qi, Z. Jiang, Q. Yu, C. Shao, H. Zhang, H. Yue, B. Ma, Y. Wang, C. Liu, X. Meng et al., “Machine learning-based ct radiomics model for predicting hospital stay in patients with pneumonia associated with sars-cov-2 infection: A multicenter study,” medRxiv, 2020.

[26] M. Nishino, “Perinodular radiomic features to assess nodule microenvironment: Does it help to distinguish malignant versus benign lung nodules?” 2019.

[27] L. Li, L. Qin, Z. Xu, Y. Yin, X. Wang, B. Kong, J. Bai, Y. Lu, Z. Fang, Q. Song et al., “Using artificial intelligence to detect covid-19 and community-acquired pneumonia based on pulmonary ct: evaluation of the diagnostic accuracy,” Radiology, vol. 296, no. 2, 2020.

[28] H. X. Bai, R. Wang, Z. Xiong, B. Hsieh, K. Chang, K. Halsey, T. M. L. Tran, J. W. Choi, D.-C. Wang, L.-B. Shi et al., “Ai augmentation of radiologist performance in distinguishing covid-19 from pneumonia of other etiology on chest ct,” Radiology, p. 201491, 2020.

